# HIV ESTIMATION USING POPULATION-BASED SURVEYS WITH NON-RESPONSE: A PARTIAL IDENTIFICATION APPROACH^*^

**DOI:** 10.1101/2023.06.03.23290936

**Authors:** Oyelola A. Adegboye, Tomoki Fujii, Denis Heng-Yan Leung, Li Siyu

**Author notes:** Citation: Adegboye OA, Fujii T, Leung DH-Y, Siyu L. HIV estimation using population-based surveys with non-response: a partial identification approach. Pages. DOI:000000/11111.

## Abstract

**Background:** HIV estimation using data from the Demographic and Health Surveys (DHS) is limited by the presence of non-response and test refusals. Conventional adjustments such as imputation require the data to be missing at random. Methods that use instrumental variables allow the possibility that prevalence is different between the respondents and non-respondents, but their performance depends critically on the validity of the instrument.

**Methods:** Using Manski’s partial identification approach, we form instrumental variable bounds for HIV prevalence from a pool of candidate instruments. Our method does not require all candidate instruments to be valid. We use a simulation study to evaluate our method and compare it against its competitors. We illustrate the proposed method using DHS data from Zambia.

**Results:** Our simulations show that imputation leads to seriously biased results even under mild violations of non-random missingness. Using worst case identification bounds that do not make assumptions about the non-response mechanism is robust but not informative. By taking the union of instrumental variable bounds balances informativeness of the bounds and robustness to inclusion of some invalid instruments.

**Conclusions:** Non-response and refusals are ubiquitous in population based HIV data such as those collected under the DHS. Partial identification bounds provide a robust solution to HIV prevalence estimation without strong assumptions. Union bounds are significantly more informative than the worst case bounds, without sacrificing credibility.

**Key messages:**

- Partial identification bounds are useful for HIV estimation when data are subject to non-response bias
- Instrumental variables can narrow the width of the bounds but validity of an instrument variable is an untestable hypothesis
- This paper proposes pooling candidate instruments and creating union bounds from the pool
- Our approach significantly reduces the width of the worst case bounds without sacrificing robustness

## 1 Background

In sub-Saharan Africa, home to around 23 million people living with HIV [1], accurate measurement of the trends of important diseases such as HIV is essential for governments to design policies and aid programs. In the past two decades, national population-based surveys from the Demographic and Health Survey (DHS) system have become an important source for such measurement [2, 3]. A major challenge in using these data is the potential bias from missing data created by non-response. There is much evidence that the non-respondents may have patterns of outcome and/or behaviour that are very different from those of the rest of the population [4, 5].

One reason why non-response has garnered significant attention from researchers is the complexity of the problem [6]. Non-response is not a result of a single source or a well-defined situation, as it is widely recognized. Instead, the causes and processes that lead to non-response are diverse and often depend on multiple factors, including the surveyed population, the outcome’s nature, and the survey’s design and implementation. The most challenging aspect of this problem is that information about non-respondents is typically limited, making it challenging for surveyors to determine the reason behind a non-response [6] In the context of HIV survey, non-response arises primarily from two sources– non-contacts and refusals. The processes leading to these two types of non-responses are believed to be distinct. But for ease of discussion, we use these terms interchangeably. We return to distinguish them in the empirical study.

A primary concern when reporting HIV prevalence estimates using DHS data is potential bias resulting from non-response. Some relevant earlier works on non-response bias in HIV estimation using data from the DHS system include [3] and [7], who carried out multi-country surveys of response rates and evaluated non-response bias. [4] examined non-response bias in a nine-country study. They assumed non-response is non-informative and estimated the prevalence among the non-respondents by multiple imputation. Similarly, [5] used a logistic regression to predict the HIV prevalence among the non-respondents under a non-informative non-response assumption in a twelve-country study. [8] and [9] corrected refusal bias in population surveys by using auxiliary longitudinal data. Their method relies on the assumption that the refusal behaviour in different populations are comparable. [10, 11, 12] adjusted non-response bias by a Heckman-type selection model [13], which allows non-response to be informative but requires the existence of a valid instrumental variable that satisfies the exclusion criteria of explaining non-response but not the outcome. [14] constructed bounds based on the partial identification approach of [15, 16]. Under this approach, the unknown quantity of interest can only be identified to within a set of bounds, whose width depends on the knowledge, or lack thereof, about the missing data. In this sense, the bounds are “worst case” bounds since no assumptions are made regarding the missingness process. Worst case bounds are often considered overly conservative in practice. [14] used restrictions implied by the dynamics of HIV (ie., an infected person remains infected over time while an uninfected person cannot be infected earlier) and instrumental variables to narrow the width of the identification region.

Methods that use instrumental variables allow the possibility that HIV prevalence is different between the respondents and those who refuse testing. However, valid instruments about the non-response mechanism are notoriously difficult to find. Furthermore, whether an instrument is valid is not a testable hypothesis. This paper aims to solve this conundrum. We espouse the view that, due to missing data, a study with missing data can never achieve as much as it would have had there been no missing data. This view departs from the conventional wisdom that, with sufficient assumptions and modelling, that a study with missing data can be restored to the state as if there were no missing data, save the fewer observations. Under the conventional perspective, unknown quantities of interest can be estimated using point estimates, or “point identified”, with an adjustment to the reduced information, and then inferential tools such as confidence intervals and hypothesis tests can be carried out as usual. In our view the uncertainty created by the missing data and our inability in pinpointing the exact causes of missingness must be embedded into the formulation of the analysis strategy.

Theoretically, if we do not know whether an instrument is valid, we can take multiple candidate instruments. Indeed, in observational epidemiological studies that are subject to confounding or reverse causation bias, the use of genetic variants as proxies for environmentally modifiable exposures may lead to a hundred or more candidate instruments [17]. However, among the instruments under consideration, we do not know which ones are valid. We propose a two-stage modification of Manski’s partial identification approach to solve this problem. Assume, *s > a*, where *a* is the minimum number of valid instruments out of the *L* candidate instruments under consideration. For each candidate, we can use Manski’s approach to form bounds. Then even though we do not know the validity of individual instruments, the union of bounds using any set of *L − a* + 1 individual candidates is guaranteed to correctly identify the quantity of interest.

Following [18, 19, 20] that the intersection of bounds is non-empty for any set of valid instruments to eliminate the candidates whose bounds fail to overlap with the bounds of the majority of the candidates, we then take the intersection of the union bounds from all possible sets of *L − a* + 1 instruments to form a new set of bounds. This step substantially narrows the bounds in some cases without sacrificing robustness. We carry out a simulation experiment to evaluate the proposed method. We then illustrate our method using data from the Zambia Demographic Health Surveys.

## 2 Method

We assume for each individual in the population of interest, an outcome variable *Y* is measurable and bounded. Suppose we are interested in the population mean of *Y*, E(*Y*). In general we may also be interested in E(*Y |X*) for some covariates *X*, but for brevity, we focus our discussion in the next two sections on estimating E(*Y*) since the treatment for the case with covariates is similar. Suppose a random sample of *n* is drawn from the population and in this sample, *Y* is observed only in a subset of the sample. Let *D* be a binary variable such that *D* = 1 if *Y* is observed and 0 otherwise. Using the law of iterated expectations, we can write

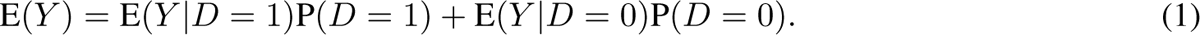

The sampling process identifies E(*Y |D* = 1), P(*D* = 1) and P(*D* = 0) = 1 *−* P(*D* = 1) but there is no information on E(*Y |D* = 0) unless we make strong assumptions about the joint distribution of *Y* and *D*. Let *K*_0_*, K*_1_ be, respectively, the lower and upper bounds of *Y*. Furthermore, write *µ ≡* E(*Y*), *µ_d·_ ≡* E(*Y |D* = *d*). The worst case partial identification bounds [15] for *µ* are

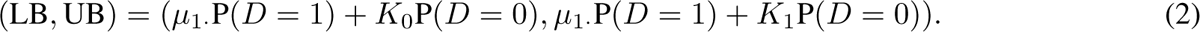

### 2.1 Bounds using instruments

The worst case bounds (2) are guaranteed to identify E(*Y*) by construction. However, they are often criticised for being too wide to be informative. The worst case bounds can be improved if additional assumptions are made. Let *V* be an instrumental variable with discrete values *v ∈ V*, such that,

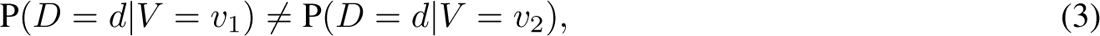

and

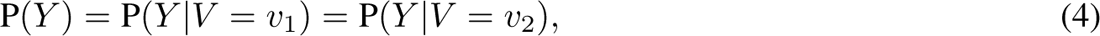

for *d* = 0, 1, all values *v*_1_*, v*_2_ *∈ V* and *v*_1_ *̸*= *v*_2_. Write *µ_·v_ ≡* E(*Y |V* = *v*) and *µ_dv_≡* E(*Y |D* = *d, V* = *v*). Since (4) implies E(*Y |V* = *v*) = E(*Y*) = *µ*, it follows that [18],

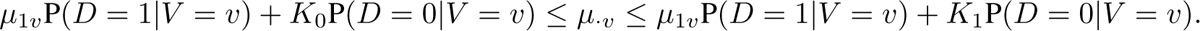

The inequalities imply

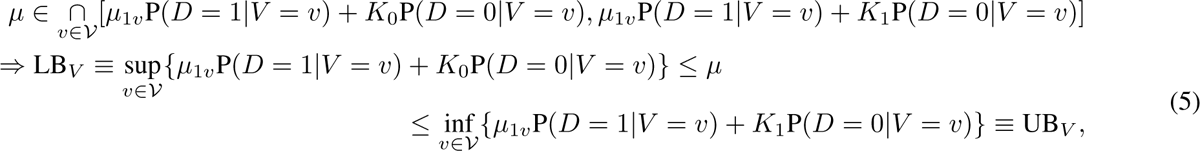

where (LB*_V_,* UB*_V_*) gives a set of IV lower and upper bounds for *µ*.

In practice, more than one instrument is usually used in a particular study [see, e.g., 17, 21]. Suppose there are *L* candidate instruments, and all we can assume is at least one of the *L* candidates is valid. Then, if some turn out to be invalid, (5) may fail to identify E(*Y*) for these instruments. To address this, suppose we create the following “union” bounds:

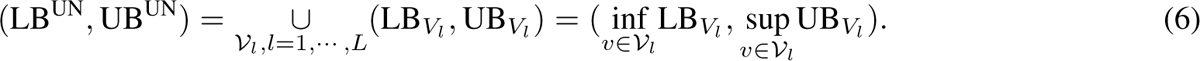

It is trivial to see that (LB^UN^, UB^UN^) identifies E(*Y*) as long as at least one of the candidate instruments is valid. However, a simple examination of (LB^UN^, UB^UN^) reveals that as *L* increases, so will the width of (LB^UN^, UB^UN^). The wider a set of bounds, the less informative it is in identifying E(*Y*). Hence it would be of interest to eliminate among the *L* instruments, those that do not contribute to the identification of E(*Y*). To continue, we assume that the true number of valid instruments, *s* is known to satisfy *s > a ≥* 1 for some known *a*. Under this assumption, each subset of (*L − a* + 1) instruments must contain at least one valid instrument. Hence, the union bounds formed by each subset is guaranteed to identify E(*Y*). For any two sets of bounds that both include *E*(*Y*), their intersection must be non-empty, and also correctly identify E(*Y*). We therefore propose to find the intersection of all union bounds formed with any (*L − a* + 1) instruments among the *L* instruments, because it will also identify E(*Y*) but be no longer than any of these union bounds.

Applying the bounds empirically incurs uncertainty and this uncertainty can be incorporated in the form of confidence intervals. A confidence interval should have a high asymptotic probability of containing both (LB, UB) or *µ*. Here, we follow the approach suggested in [22] for forming confidence intervals. Justifications and further details about the proposed bounds, and confidence intervals are given in the Supplementary materials.

### 3 Simulation study

We use a simulation study to evaluate our proposed bounds (6). We assume the response *Y* is binary. We fix the values of *s* and *L* at 3 and 5, respectively. The instruments are all binary with prevalence of 0.5 and mutually independent of each other.

We generate *Y* using a logistic model

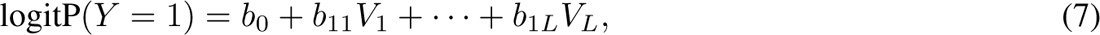

where the coefficients *b*_1_ = (*b*_11_*, · · ·, b*_1_*_L_*)*^T^* give the association between the instruments and *Y*. A non-zero value of *b*_1_*_j_* induces an association and therefore renders the instrument invalid. We use two different combinations for *b*_1_: 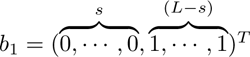 and 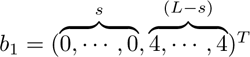. For both situations, we assume without loss of generality the first *s* instruments are valid while the remaining *L − s* are invalid. In the former, (4) is weakly violated by the invalid instruments while the violation of (4) is strong for the latter.

The non-response indicator *D* is generated using another logistic model

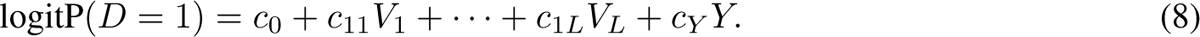

The coefficients *c*_1_ = (*c*_11_*, · · ·, c*_1_*_L_*)*^T^* give the association between each instrument and *D*. We consider two situations, (a) Strong instruments: *c*_1_ = (5*, · · ·,* 5) and (b) Strong + weak instruments: *s* coefficients are randomly given a value of 5 and the remaining *L − s* are given a value of 0.5. The coefficient *c_Y_* is used to model the association of *D* to the outcome *Y*, and hence selection bias. When *c_Y_* = 0, then there is no selection bias when conditioned on the observed covariates. We consider two choices of *c_Y_* = *−*0.1*||c*_1_*||* and *−*0.3*||c*_1_*||*, where the symbol *|| · ||* stands for the sum of the coefficients *c*_11_*, …, c*_1_*_L_*. We use negative association to reflect that in practice, we expect those who are HIV positive are less likely to have an HIV test. These two values for *c_Y_* correspond to mild to moderate selection bias. We use *c*_0_ to calibrate the average non-response rate, 1 *−* E(*D* = 1), to be 0.1 and 0.3 over the simulations.

Since *Y* is binary, the bounds for *Y* are (*K*_0_*, K*_1_) = (0, 1). Throughout the simulation study, a sample size of *n* = 1000 observations is used. We use 1000 simulation runs for each combination of parameters. Confidence intervals are approximated using the method described in the Supplementary materials. These confidence intervals require estimates of the standard errors of the bounds, which can be carried out using bootstrapping. Throughout, we use 100 bootstrap for this purpose.

A standard approach to adjust HIV prevalence estimates for survey non-response is by imputation [23]. Using imputation, the missing outcomes are imputed using predicted prevalence based on observed information such as demographic, socio-economic and behavioural variables from those who were tested. We compare this method to the partial identification method. For the imputation method, we use all the observed variables in the simulation study, i.e., the instruments. For partial identification, we used the worst case bounds that do not make any assumptions, and also the method proposed in this article.

Table 1 gives the simulation results. Each combination of parameters corresponds to four rows of results. The first row shows the proportion of times, out of 1000 simulations, the approximate 95 percent confidence intervals include E(*Y*). The second row gives the lower confidence limits, averaged over 1000 simulations. The third row gives the upper confidence limits, averaged over 1000 simulations. The fourth row gives the average width of the confidence intervals.

**Table 1:** Partial identification of E(*Y*) with *L* = 5 instruments and *s* = 3 valid instruments with E(*Y*) fixed at 0.15. Results are stratified by average non-response rate 1 *−* E(*D*) = 0.1 or 0.3; instruments either all strong or a mixture of strong + weak; the last *L − s* instruments either weakly or strongly violate (4); and mild or moderate selection bias.

| Instrument Strength | Non-response rate | Selection bias |  | Imputation | Worst case bounds | IV bounds |
| --- | --- | --- | --- | --- | --- | --- |
| Strong | 0.1 | Mild | Coverage | 0.985 | 1 | 0.972 |
|  |  |  | Lower CI | 0.119 | 0.114 | 0.123 |
|  |  |  | Upper CI | 0.162 | 0.262 | 0.218 |
|  |  |  | Width | 0.043 | 0.148 | 0.095 |
| Strong + Weak | 0.1 | Mild | Coverage | 1 | 1 | 0.868 |
|  |  |  | Lower CI | 0.125 | 0.121 | 0.127 |
|  |  |  | Upper CI | 0.169 | 0.266 | 0.248 |
|  |  |  | Width | 0.044 | 0.145 | 0.121 |
| Strong | 0.3 | Mild | Coverage | 0.075 | 1 | 1 |
|  |  |  | Lower CI | 0.095 | 0.084 | 0.108 |
|  |  |  | Upper CI | 0.137 | 0.434 | 0.323 |
|  |  |  | Width | 0.042 | 0.35 | 0.216 |
| Strong + Weak | 0.3 | Mild | Coverage | 0.078 | 1 | 1 |
|  |  |  | Lower CI | 0.088 | 0.072 | 0.098 |
|  |  |  | Upper CI | 0.134 | 0.42 | 0.287 |
|  |  |  | Width | 0.045 | 0.349 | 0.189 |
| Strong | 0.1 | Moderate | Coverage | 0.003 | 1 | 1 |
|  |  |  | Lower CI | 0.091 | 0.088 | 0.11 |
|  |  |  | Upper CI | 0.129 | 0.234 | 0.201 |
|  |  |  | Width | 0.039 | 0.146 | 0.092 |
| Strong + Weak | 0.1 | Moderate | Coverage | 0.118 | 1 | 1 |
|  |  |  | Lower CI | 0.102 | 0.098 | 0.116 |
|  |  |  | Upper CI | 0.142 | 0.239 | 0.207 |
|  |  |  | Width | 0.04 | 0.141 | 0.091 |
| Strong | 0.3 | Moderate | Coverage | 0 | 1 | 1 |
|  |  |  | Lower CI | 0.05 | 0.046 | 0.07 |
|  |  |  | Upper CI | 0.081 | 0.388 | 0.287 |
|  |  |  | Width | 0.031 | 0.342 | 0.218 |
| Strong + Weak | 0.3 | Moderate | Coverage | 0 | 1 | 1 |
|  |  |  | Lower CI | 0.065 | 0.058 | 0.083 |
|  |  |  | Upper CI | 0.1 | 0.399 | 0.313 |
|  |  |  | Width | 0.035 | 0.341 | 0.23 |

When non-response probability is 0.1 and selection bias is mild, 95% confidence intervals using all three methods have high probabilities of capturing E(*Y*). Using imputation naturally leads to much narrower confidence intervals. Between the partial identification bounds, the IV bounds proposed in this paper produces much narrower confidence interval but at the expense of not capturing E(*Y*) in finite samples.

In all other situations, using imputation leads to grossly biased confidence intervals that fail to capture E(*Y*) in almost all simulation runs. Recall that E(*Y*) is calibrated to be at 0.15 in all simulations so the imputation confidence intervals under estimate the true prevalence. The advantage of the IV bounds confidence intervals over the worst case confidence intervals mirror those when non-response probability is 0.1 and selection bias is mild. Additional simulations have been carried out. The results are given in the Supplementary materials. The conclusion from the additional simulations is similar to those presented here.

### 4 HIV prevalence in Zambia

The primary data source for this study is the 2007 Zambia DHS. The 2007 Zambia DHS is the fourth survey in the Zambia DHS series and provides population-level health estimates, including data useful in monitoring and evaluating population, health, and nutrition programs.

A total of 7969 households were selected for the 2007 Zambia DHS, of which 7326 were occupied. The shortfall was largely due to households that were away for an extended period of time and structures that were found to be vacant at the time of the interview. Of the occupied households a total of 7146 were successfully interviewed. The interviews collected basic demographic information (e.g., age, sex), socio-economic status (e.g., educational attainment) as well as basic household characteristics (e.g., household possessions and dwelling characteristics).

In the interviewed households, 7406 females were eligible for interview and HIV testing, while the number of males was 7146. The individual interviews collected information such as work and background characteristic, marriage and sexual activities, and awareness and attitudes towards HIV. In the women’s interviews, additional questions about reproductive history and child heath and nutrition were asked.

Of the women and men eligible for individual interviews, 1695 (22.8%) of the women and 1983 (27.8%) of the men refused or did not complete an HIV test. The primary reason for non-response among eligible men was the failure to find individuals at home despite repeated visits to the household, followed by refusal to be interviewed. The substantially lower response rate for men reflects the more frequent and longer absence of men from the households.

The interviews in the 2007 Zambia DHS were carried out by 12 teams made up of 12 supervisors, 12 editors, 36 female interviewers, and 36 male interviewers. Each team consisted of one supervisor, one female field editor, one laboratory technician, three female interviewers, and three male interviewers. The interviews and questionnaires were translated from English into one of seven major local language groups: Nyanja, Bemba, Kaonde, Lunda, Lozi, Tonga, and Luvale.

The observed prevalence of HIV positive among the cases with results, stratified by age, are given in Table 2. Even though in this study, the proportions of non-response is modest, we shall see that using instruments still bring improvements on inferences in some cases.

**Table 2:**
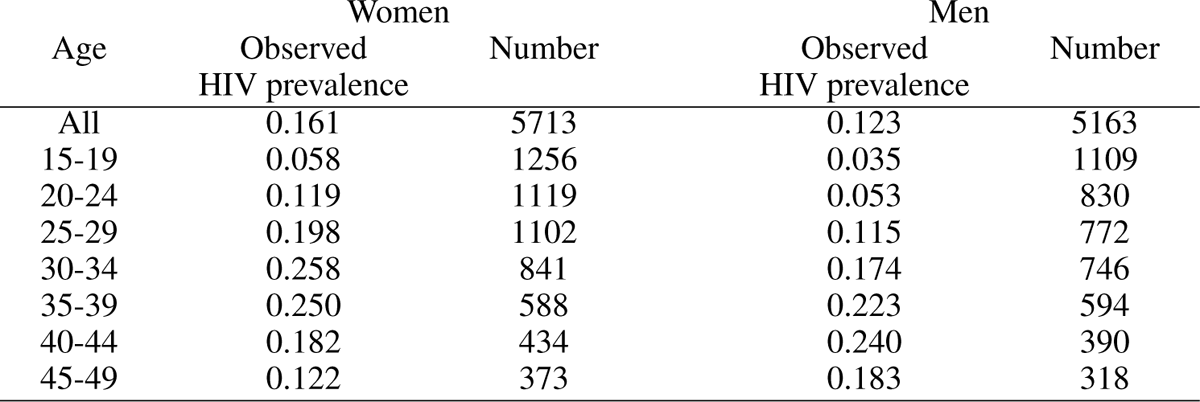
Observed proportions of HIV positive among the tested in 2007 Zambia DHS

We examine HIV prevalence between genders, and across different age groups. Previous studies have suggested that variables related to the data collection process may be used as instruments because they affect the response probability but are unlikely to have a direct effect on the outcome [12, 24]. For example, an experienced interviewer or an interviewer of a similar age as the interviewee may have a better chance of eliciting a positive response. Furthermore, whether the language of the interview or questionnaire is the same language as the interviewee may affect response rate. It has also been argued that timing of the first interview attempt that coincides with the economic cycle affects the probability of finding the interviewees at home. Individuals selected to be interviewed on the first day of the interviews within a cluster of households will also have more chances to be contacted even if they are not at home, giving rise to a higher response probability. Finally we also consider a variable based on the individual’s attitude to HIV. The current literature finds that more negative attitudes are associated with refusal of an HIV test or never having had an HIV test in sub-Saharan Africa [25, 26].

The final list of instrumental variables we use are: iv.lan (whether the language used in the questionnaire or interview is the same as the respondent’s language, yes *vs.* no), iv.firstday (whether the interview was conducted on the first day of the interviews, yes *vs.* no), iv.interviewer (number of interviews the interviewer has performed, *<* 50, 50 *−* 100, 100 *−* 200, *>* 200), iv.mon (whether the interview was carried out during a month of harvest or planting, yes *vs.* no), iv.doa (whether the respondent knows someone who has died of AIDS, yes *vs.* no).

It is well known that the validity of an instrument (4) is an untestable hypothesis. Nevertheless, we can determine whether an instrument is strong by evaluating (3). Table 3 shows chi-square tests between non-response and the candidate instrumental variables we consider; all tests are highly significant.

**Table 3:** Relationship between HIV testing and some possible instrument variables

(a): Women
| D | 0 |  | 1 |  | Test |
| --- | --- | --- | --- | --- | --- |
| Variable | N | Percent | N | Percent |  |
| iv.lan | 1695 | | 5713 | | $\chi^2 = 24.706^{***}$ |
| ... 0 | 727 | 42.9% | 2845 | 49.8% |  |
| ... 1 | 968 | 57.1% | 2868 | 50.2% |  |
| iv.firstday | 1695 | | 5713 | | $\chi^2 = 22.774^{***}$ |
| ... 0 | 1094 | 64.5% | 3315 | 58% |  |
| ... 1 | 601 | 35.5% | 2398 | 42% |  |
| iv.interviewer | 1433 | | 5713 | | $\chi^2 = 28.929^{***}$ |
| ... 0 | 74 | 5.2% | 256 | 4.5% |  |
| ... 1 | 190 | 13.3% | 807 | 14.1% |  |
| ... 2 | 695 | 48.5% | 2369 | 41.5% |  |
| ... 3 | 474 | 33.1% | 2281 | 39.9% |  |
| iv.mon | 1695 | | 5713 | | $\chi^2 = 82.634^{***}$ |
| ... 0 | 311 | 18.3% | 1688 | 29.5% |  |
| ... 1 | 1384 | 81.7% | 4025 | 70.5% |  |
| iv.doa | 1695 | | 5713 | | $\chi^2 = 14.097^{***}$ |
| ... 0 | 688 | 40.6% | 2616 | 45.8% |  |
| ... 1 | 1007 | 59.4% | 3097 | 54.2% |  |

Statistical significance markers: \* p<0.1; \*\* p<0.05; \*\*\* p<0.01 (b): Men
| D | 0 |  | 1 |  | Test |
| --- | --- | --- | --- | --- | --- |
| Variable | N | Percent | N | Percent |  |
| iv.lan | 1983 | | 5163 | | $\chi^2 = 160.186^{***}$ |
| ... 0 | 739 | 37.3% | 2789 | 54% |  |
| ... 1 | 1244 | 62.7% | 2374 | 46% |  |
| iv.firstday | 1983 | | 5163 | | $\chi^2 = 4.267^{**}$ |
| ... 0 | 1251 | 63.1% | 3118 | 60.4% |  |
| ... 1 | 732 | 36.9% | 2045 | 39.6% |  |
| iv.interviewer | 1339 | | 5161 | | $\chi^2 = 13.453^{***}$ |
| ... 0 | 167 | 12.5% | 583 | 11.3% |  |
| ... 1 | 62 | 4.6% | 373 | 7.2% |  |
| ... 2 | 493 | 36.8% | 1791 | 34.7% |  |
| ... 3 | 617 | 46.1% | 2414 | 46.8% |  |
| iv.mon | 1983 | | 5163 | | $\chi^2 = 200.984^{***}$ |
| ... 0 | 293 | 14.8% | 1621 | 31.4% |  |
| ... 1 | 1690 | 85.2% | 3542 | 68.6% |  |
| iv.doa | 1983 | | 5163 | | $\chi^2 = 60.931^{***}$ |
| ... 0 | 650 | 32.8% | 2216 | 42.9% |  |
| ... 1 | 1333 | 67.2% | 2947 | 57.1% |  |

Statistical significance markers: \* p<0.1; \*\* p<0.05; \*\*\* p<0.01

We assume *a* = 3, that is, at least 3 out of the 5 candidates are valid. In any survey such as the 2007 Zambia DHS, non-response, and the potential for an associated bias, is always a concern. The standard procedure is an imputation analysis on those who are not tested to adjust for potential biases [27]. The individuals in the survey can be classified into one of three groups: (a) those who participated in the household and individual surveys and tested (b) those who participated in the household and individual surveys but not tested and (c) those who only participated in the household surveys. For those in groups (b) and (c), their HIV test results are absent.

For individuals in groups (b) and (c), their probability of HIV is predicted based on multivariate models using data from those who were tested. A logistic regression model is used to calculate HIV probability separately for groups (b) and (c). For group (b), the variables used in the model include the following household survey variables: age, education, wealth quintile, residence, and geographic region, as well as the following variables from the individual survey: marital union, current work status, media exposure, religion, sexually transmitted infections (STIs) or STI symptoms in past 12 months, cigarette smoking/tobacco use, age at first sex, number of sex partners in past 12 months, higher-risk sex in past 12 months, condom use at last sex in past 12 months, and willingness to care for a family member with AIDS. Prediction for group (c) uses only the household variables. The models are used to impute HIV statuses for individuals in groups (b) and (c) and the results are combined with those in group (a) to form adjusted HIV prevalence estimates for the population.

For all estimates, the data are weighted by survey weights. For individuals in group (a), HIV weights were used, for individuals in group (b), the individual survey weights were used and for those in group (c), household survey weights were used.

We compare adjustments using standard imputation with those using partial identification bounds. For partial identification bounds, we report results based on the worst case bounds as well as the instrumental variable bounds. For brevity, we only report the 95% confidence intervals (CIs) in Table 4a-b. We also include confidence intervals based on the observed, unadjusted prevalence among the tested individuals. The results are stratified by gender and by age groups.

**Table 4:** 95% confidence intervals for HIV prevalence estimates in 2007 Zambia DHS

|  |  | (a) Women |  |  |  | (b) Men |  |  |  |
| --- | --- | --- | --- | --- | --- | --- | --- | --- | --- |
| Age |  | Unadjusted | Imputation | Worst case bounds | IV bounds | Unadjusted | Imputation | Worst case bounds | IV bounds |
| All | LCI | 0.153 | 0.153 | 0.118 | 0.123 | 0.116 | 0.118 | 0.084 | 0.087 |
|  | UCI | 0.169 | 0.169 | 0.361 | 0.339 | 0.13 | 0.133 | 0.375 | 0.322 |
|  | Width | 0.016 | 0.016 | 0.243 | 0.216 | 0.014 | 0.015 | 0.291 | 0.235 |
| 15-19 | LCI | 0.038 | 0.041 | 0.029 | 0.023 | 0.018 | 0.021 | 0.014 | 0.011 |
|  | UCI | 0.078 | 0.073 | 0.305 | 0.274 | 0.051 | 0.045 | 0.32 | 0.278 |
|  | Width | 0.04 | 0.032 | 0.276 | 0.251 | 0.033 | 0.024 | 0.306 | 0.267 |
| 20-24 | LCI | 0.099 | 0.105 | 0.073 | 0.076 | 0.034 | 0.039 | 0.024 | 0.025 |
|  | UCI | 0.139 | 0.137 | 0.36 | 0.331 | 0.073 | 0.067 | 0.352 | 0.317 |
|  | Width | 0.04 | 0.032 | 0.287 | 0.255 | 0.039 | 0.028 | 0.328 | 0.292 |
| 25-29 | LCI | 0.181 | 0.185 | 0.142 | 0.147 | 0.097 | 0.105 | 0.068 | 0.085 |
|  | UCI | 0.216 | 0.213 | 0.392 | 0.383 | 0.132 | 0.129 | 0.406 | 0.358 |
|  | Width | 0.035 | 0.028 | 0.25 | 0.236 | 0.035 | 0.024 | 0.338 | 0.273 |
| 30-34 | LCI | 0.237 | 0.239 | 0.187 | 0.202 | 0.156 | 0.171 | 0.112 | 0.124 |
|  | UCI | 0.279 | 0.272 | 0.44 | 0.401 | 0.192 | 0.197 | 0.425 | 0.386 |
|  | Width | 0.042 | 0.033 | 0.253 | 0.199 | 0.036 | 0.026 | 0.313 | 0.262 |
| 35-39 | LCI | 0.223 | 0.236 | 0.174 | 0.178 | 0.199 | 0.211 | 0.145 | 0.154 |
|  | UCI | 0.278 | 0.278 | 0.443 | 0.408 | 0.246 | 0.246 | 0.459 | 0.378 |
|  | Width | 0.055 | 0.042 | 0.269 | 0.23 | 0.047 | 0.035 | 0.314 | 0.224 |
| 40-44 | LCI | 0.151 | 0.163 | 0.12 | 0.131 | 0.212 | 0.222 | 0.163 | 0.168 |
|  | UCI | 0.212 | 0.211 | 0.391 | 0.342 | 0.268 | 0.262 | 0.455 | 0.413 |
|  | Width | 0.061 | 0.048 | 0.271 | 0.211 | 0.056 | 0.04 | 0.292 | 0.245 |
| 45-49 | LCI | 0.09 | 0.1 | 0.072 | 0.073 | 0.153 | 0.17 | 0.116 | 0.129 |
|  | UCI | 0.155 | 0.151 | 0.333 | 0.315 | 0.211 | 0.214 | 0.431 | 0.382 |
|  | Width | 0.065 | 0.051 | 0.261 | 0.242 | 0.058 | 0.044 | 0.315 | 0.253 |

We hereafter focus the discussion on men’s results, the women’s results exhibit similar patterns. The imputation method uses models based on data from the tested, and hence implicitly it assumes that conditioned on the covariates used in the models, a non-tested individual has the same propensity of HIV as a tested individual. This fact is borne out in the 95% CIs using imputation. All of them include the corresponding observed prevalence among the tested in Table 2 and they are all very similar to the corresponding CIs based on the unadjusted prevalence estimates. Each imputation CI is never wider than its unadjusted counterpart due to the additional observations used. Both have relatively short widths due to the large sample sizes in this study.

Using partial identification, the corresponding CIs are much wider than those using imputation. The much wider CIs using partial identification reflect the uncertainties we have about the actual HIV status for the non-tested individuals. The lower limits of the partial identification CIs are also in general quite a bit smaller than the corresponding imputation lower limit. The reason is that a lower partial identification bound is derived by assuming all non-tested individuals are HIV negative (for a given value of instrument for the IV bound), whereas imputation assumes the non-tested are the same as the tested, given the covariates. Similarly, the upper limits of the partial identification CIs are much higher than those given by imputation, since the upper partial identification bounds result from assuming all non-tested are HIV positive (for a given value of instrument for the IV bound).

The most significant difference between the partial identification CIs and the imputation CIs lie in those situations where the observed prevalence among the tested is low. For example, in the male aged 15-19 group. The upper limit of the imputation CI is 0.045, against the upper limits of 0.32 and 0.278, respectively, for the worst case and instrumental variable partial identification CIs. The reason for the very low upper limit for the imputation CI is that it assumes those non-tested also have similarly low prevalence as the tested individuals. In contrast, the partial identification approach allows for the possibility that even if a moderate proportion of the non-tested are actually HIV positive, the prevalence would change significantly upwards.

Between the two partial identification methods, the worst case scenario makes no assumptions and the resulting CIs are wider than those derived using the instrumental approach proposed in this paper. Since the width of a CI gives its precision, the method proposed here is always more precise than the worst case CIs. In some cases, such as males aged 35-39, the gain in precision approaches 30%. Two other observations are worth noting. First, as expected, all CIs using the proposed method have a larger lower confidence limit that the corresponding worst case CIs. A second observation is the narrowing in widths in the CIs in the proposed method mainly comes from a much smaller upper limit than the corresponding worst case CIs. This advantage is brought about when the population is stratified by different levels of a valid instrument. if the proportion of tested individuals is higher at a particular level, the more precise information from such a group can be used to infer about HIV prevalence of the entire population.

## 5 Discussion

Existing studies on refusal bias in the estimation of HIV prevalence typically either provide some evidence of the existence of the bias or try to correct for the bias by making some (often strong) behavioural assumptions about the subjects. In this paper, we have instead derived plausible lower and upper bounds for HIV prevalence under mild and intuitive assumptions. This approach is potentially useful because it is often difficult to validate or falsify an underlying assumption. Furthermore, it shows that a carefully designed and implemented localised study may also be helpful for understanding the magnitude of non-response bias.

Partial identification approach using instruments has been widely used in the fields of Social Sciences and Economics, though rare in Epidemiology and Public Health. As with other methods that exploits instruments, the key to the success of this approach is the validity of the instruments used to create the bounds. However, it is well known that the exclusion restriction assumption is a non-testable hypothesis. This paper offers a novel and simple solution to this challenge by taking multiple candidate instruments. If at least one instrument in the pool of candidates is valid, the proposed approach creates bounds that, in large samples, identify the true prevalence. The approach offered in this paper is especially useful for practitioners because normally there are multiple variables, eg., interviewing process, interviewer characteristics, etc., that are candidates to be considered as instruments and yet there is no way to determine which one(s) is(are) valid. Using a large pool increases the chance of finding at least one that is valid but at the same time, induces the possibility of including invalid ones. The proposed method solves this conundrum.

Our proposed method is similar to that proposed in [28] for estimating causal effects when some instruments are possibly invalid. [28] also considers a union method but their context and process are different from the present paper. In their paper, the goal is to obtain a confidence interval of some causal effect. They also assume a pool of *N* instruments with no more than *s^∗^* valid (in our notations). For each set of *s^∗^ −* 1 instruments, they form confidence interval of the causal effect. They then take the union of confidence intervals over all 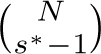 sets of instruments. On the contrary, our method first creates the partial identification bound using each instrument, then find the union of bounds from every set of *N − s^∗^* + 1 instruments. In [28], the interval is narrowed by pretesting and eliminating possibly invalid instruments. In our paper, no tests are used, instead, we take the intersection of the 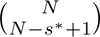 union bounds.

There are two cautionary notes to be made about the proposed method. The first is that it is advisable not to include too many instruments, particularly highly dubious ones. The goal of the proposed method is to create bounds that identify the prevalence robust to invalid instruments. However, if we add mostly invalid instruments, *L* will increase without corresponding increase in *a*. This will result in the increase in the number of instruments, *L − a* + 1, leading to a wide confidence bound in the proposed method. Therefore, a balance must be struck as to how many and what instruments should be included as candidates. Obviously, we should include as many as needed so we have comfort that some among the pool of candidates would be valid. Our simulations and empirical example suggest that just a few candidate instruments would suffice.

Second, the proposed method is not immune to the problem of weak instrumental variables. A weak instrumental variable is one that is not informative about the non-response process. Instrumental variable bounds based on weak instruments may be very wide and not much different from the worst case bounds. Therefore, we must also be judicious about the choice of instruments. Fortunately, the strength of an instrument is a testable hypothesis. We demonstrated a way this can be done in the empirical study (Table 3).

In conclusion, the proposed approach is useful for providing HIV prevalence estimates in population-based surveys where non-response is a ubiquitous phenomenon and little is known about the causes of the non-response.

## Data Availability

This study is based on publicly available data, the Zambia DHS obtained from www.dhsprogram.com. The program used in this simulation study can be obtained from https://github.com/oyeadegboye/partialindentification

## Acknowledgement

We acknowledge ORC Macro for granting us access to the Zambia DHS data.

## Conflict of Interest

None declared

## Ethics approval

This study is based on publicly available data, therefore ethical approval is not required.

## Author contributions

O.A.A., T.F., D.H-Y.L. and L.S. all drafted the manuscript. D.H-Y.L. and L.S. conducted simulation analyses. A.F.-S.O.A.A., T.F., D.H-Y.L. and L.S. conducted data analyses. All authors contributed to the design of the study. All authors critically reviewed and revised the manuscript.

## Supplementary material

In this supplement, we provide results for additional materials and clarifications.

### S.1 Bounds using instruments

We assume for each individual in the population of interest, an outcome variable *Y* is measurable. Suppose we are interested in the population mean of *Y*, E(*Y*). In general we may also be interested in E(*Y |X*) for some covariates *X*, but for brevity, we focus our discussion in the next two sections on estimating E(*Y*) since the treatment for the case with covariates is similar. Suppose a random sample of *n* is drawn from the population and in this sample, *Y* is observed only in a subset of the sample. Let *D* be a binary variable such that *D* = 1 if *Y* is observed and 0 otherwise. Using the law of iterated expectations, we can write

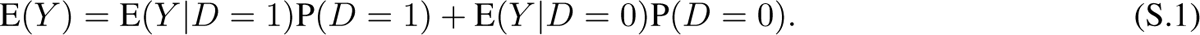

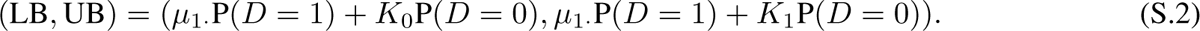

The worst case bounds (S.2) are guaranteed to identify E(*Y*) by construction. However, they are often criticised for being too wide to be informative. The worst case bounds can be improved if additional assumptions are made. Let *V* be an instrumental variable with discrete values *v ∈ V*, such that,

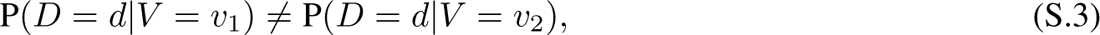

and

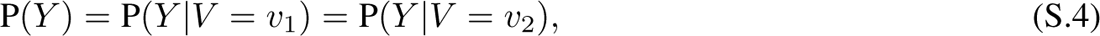

for *d* = 0, 1, all values *v*_1_*, v*_2_ *∈ V* and *v*_1_ *̸*= *v*_2_. Write *µ_·v_ ≡* E(*Y |V* = *v*) and *µ_dv_ ≡* E(*Y |D* = *d, V* = *v*). Since (S.4) implies E(*Y |V* = *v*) = E(*Y*) = *µ*, it follows that [18], *∀v ∈ V*,

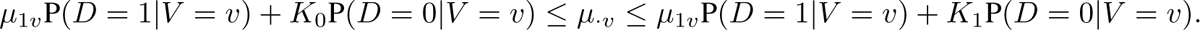

The inequalities imply

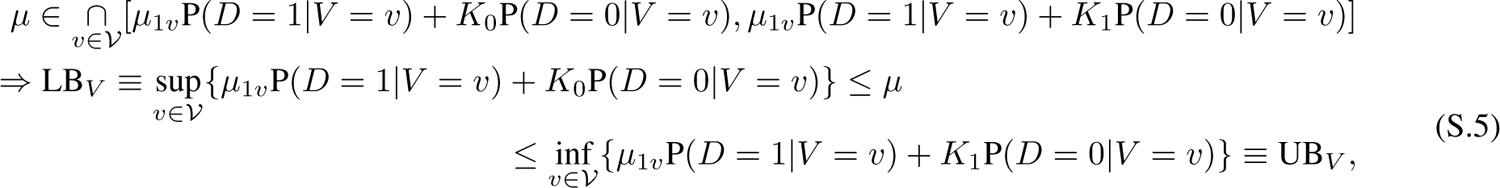

where (LB*_V_,* UB*_V_*) gives a set of IV lower and upper bounds for *µ*. It is straightforward to see that the IV bounds are guaranteed to lie within the worst case bounds, hence if *V* is observed for all individuals in the sample, a set of tighter bounds than those given by the worst case bounds can be achieved. Notice that in order for the IV bounds to work, assumptions (S.3) and (S.4) must both be satisfied. Assumption (S.4) is a necessary condition; violation of (S.4) gives an invalid instrument, which may lead to bounds that fail to identify the quantity of interest. Violation of assumption (S.3) gives a weak instrument [29]. While using a weak instrument does not lead to invalid inferences, the bounds (S.5) become uninformative. To see this last point, suppose (S.4) is satisfied but (S.3) is not, such that P(*D* = *d|V* = *v*) = P(*D* = *d*) for all *v ∈ V*; then the left hand of the inequality (S.5) becomes

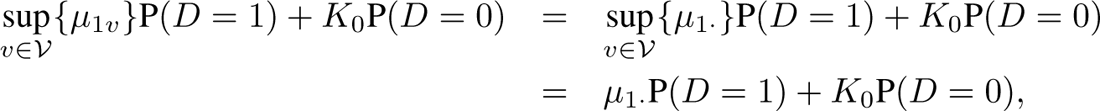

which is identical to the lower worst case bound (S.2). Similarly, the right hand side of (S.5) becomes the upper worst case bound. The observed data on *D*, however, allow us to verify whether an instrument is weak via (S.3).

In practice, more than one instrument is usually used in a particular study [see, e.g., 17, 21]. Suppose *l* candidate instruments are considered for reducing the width of the worst case bounds. Define *{V*_1_*, · · ·, V_t_}* for any arbitrary set of *t ≥* 1 instruments. Suppose there are *t* = *L >* 1 instruments such that *V_l_, l* = 1*, · · ·, L* all satisfy (S.3) and (S.4). Write for *V_l_*, the bounds (LB*_Vl_,* UB*_Vl_*). Then *µ* must also lie in the “intersection” of the bounds [19]:

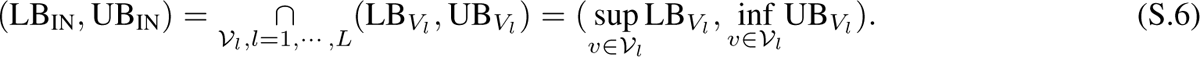

Even though the IV and intersection bounds provide refinements on the worst case bounds, these refinements are achieved at the expense of having to identify instrumental variables that satisfy assumptions (S.3) together with (S.4). It is well known that valid and informative instruments are difficult to find. More importantly, assumption (S.4) is not verifiable, and hence in practice, these bounds are anchored on our beliefs that the assumptions are satisfied. If even one of the *L* instruments is invalid, the bounds would fail to identify E(*Y*). This problem where some of the instruments may be invalid is well known in the casual inference literature. Our remedy is to create union bounds:

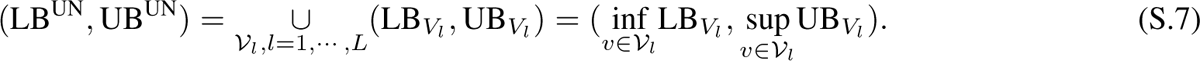

It is trivial to see that (LB^UN^, UB^UN^) identifies E(*Y*) as long as at least one of the candidate instruments is valid. To reduce the width of the union bounds, we make the assumption that the true number of valid instruments, *s* is known to satisfy *s > a ≥* 1 for some known *a*. Under this assumption, each subset of (*L − a* + 1) instruments must contain at least one valid instrument. Hence, the union bound formed by each subset is guaranteed to identify E(*Y*). For any two sets of bounds that both include *E*(*Y*), their intersection must be non-empty, and also correctly identify E(*Y*). We therefore propose to find the intersection of all union bounds formed with any (*L − a* + 1) instruments among the *L* instruments, because it will also identify E(*Y*) but be no longer than any of these union bounds.

### S.2 Confidence intervals

Applying the bounds empirically incurs uncertainty and this uncertainty can be incorporated in the form of confidence intervals. Let (LB, UB) denote a set of generic theoretical lower and upper IV bounds for *µ*. Let 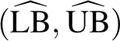 be any empirical estimate of (LB, UB). A confidence interval should have a high asymptotic probability of containing both (LB, UB) or *µ*. Here, we focus on finding an approximate *b*_0_ *×* 100 percent for *µ*. An approximate *b*_0_ *×* 100 percent confidence interval for (LB, UB) is simply of the form 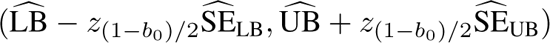, where *z*_(1_*_−b_*_0)_*_/_*_2_ is the upper (1 *− b*_0_)*/*2 *×* 100 percentile of the standard normal distribution, SE represents standard error and SE its sample analogue. As pointed out by [22], this interval would be too wide for *µ*. In fact, since (LB, UB) is a set of bounds and if we are interested in *µ*, then it will be nearer to one of 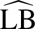 or 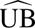 but not both simultaneously. Hence, they suggested the following bounds^†^:

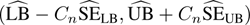

such that *C_n_* is determined by

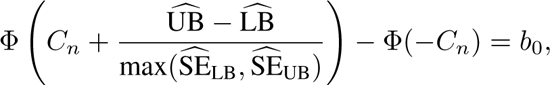

where Φ is the standard normal CDF. For example, if *b*_0_ is 0.95 such that we are interested in approximate 95% confidence intervals, then the value of *C_n_* approaches 1.64 when 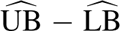 is large and it approaches 1.96 when 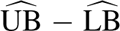 is near zero. Since 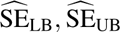 are extremely difficult to find analytically in all practical cases, following [30], we resort to bootstrapping. We sample with replacement from the data and we denote a generic bootstrap sample (*d^∗^, v^∗^, · · ·, v^∗^, y^∗^ · d^∗^*), where *i* = 1*, · · ·, n* is the index for individuals. Using each bootstrap sample, we find 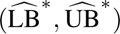 and from *B* bootstrap samples, we obtain 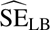 and 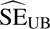.

In this section we describe additional simulation results. We use a similar set-up of the simulation study in the main paper. We assume the response *Y* is binary. We fix the values of *s* and *L* at 3 and 5, respectively. We only consider binary 0-1 instruments; The valid instruments are generated by a multivariate binary distribution, *MV B*(*µ_s_,* Σ*_s×s_*) with *µ_s_* = 0.5 *×* 1*_s_* and Σ*_s×s_* = (*σ_jj_′*)*, j, j^′^* = 1*, · · ·, s*, where *σ_jj_* = 1 and for *ρ*_1_ = *σ_jj_′, j ̸*= *j^′^*, we consider two choices of *ρ*_1_: 0 and 0.3. The first choice corresponds to the situation when all valid instruments are mutually independent, while the second choice assumes a correlation of 0.3 between each pair of instruments. We do not believe a high correlation between instruments to be a realistic situation since if two instruments are highly correlated there is no reason to use both. The invalid instruments are generated independently of the valid instruments using a *MV B*(*µ_L−s_,* Σ_(_*_L−s_*_)_*_×_*_(_*_L−s_*_)_), with *µ_L−s_* = 0.5 *×* 1*_L−s_* and Σ_(_*_L−s_*_)_*_×_*_(_*_L−s_*_)_ = (*σ_jj_′*)*, j, j^′^* = 1,, (*L − s*). We also use the same two choices of 0 and 0.3 for *ρ*_2_ = *σ_jj_′, j ̸*= *j^′^* between any two invalid instruments.

We generate *Y* using a logistic model

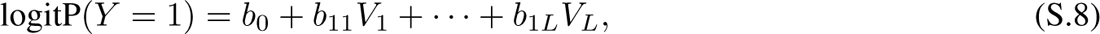

where the coefficients *b*_1_ = (*b*_11_*, · · ·, b*_1_*_L_*)*^T^* give the association between the instruments and *Y*. A non-zero value of *b*_1_*_j_* induces an association and therefore renders the instrument invalid. We use two different combinations for *b*_1_: 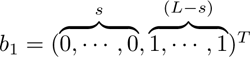; and 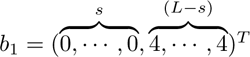. For both situations, we assume without loss of generality the first *s* instruments are valid while the remaining *L − s* are invalid. In the former, (4) is weakly violated by the invalid instruments while the violation of (4) is strong for the latter.

The non-response indicator *D* is generated using another logistic model

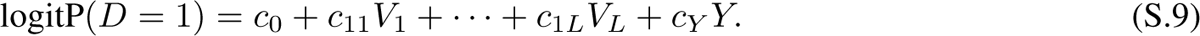

The coefficients *c*_1_ = (*c*_11_*, · · ·, c*_1_*_L_*)*^T^* give the association between each instrument and *D*. We consider two situations, (a) Strong instruments: *c*_1_ = (5*, · · ·,* 5) and (b) Strong + weak instruments: *s* coefficients are randomly given a value of 5 and the remaining *L − s* are given a value of 0.5. The coefficient *c_Y_* is used to model the association of *D* to the outcome *Y*. When *c_Y_* = 0, then there is no selection biased when conditioned on the observed covariates. We consider two choices of *c_Y_* = *−*0.1*||c*_1_*||* and *−*0.3*||c*_1_*||*, where the symbol *|| · ||* stands for the sum of the coefficients *c*_11_*, …, c*_1_*_L_*. We use negative association to reflect that in practice, we expect those who are HIV positive are less likely to have an HIV test. These two values for *c_Y_* correspond to weak to moderate associations between *Y* and *D*. We use *c*_0_ to calibrate the average non-response rate, 1 *−* E(*D* = 1), to be 0.1, 0.3, and 0.5 over the simulations.

Since *Y* is binary, the bounds for *Y* are (*K*_0_*, K*_1_) = (0, 1). Throughout the study, we use a sample size of *n* = 1000 observations for each simulation run. We use 1000 simulation runs for each combination of parameters and 100 bootstraps to estimate the standard errors of the partial identification bounds.

Tables 1(a)-(c) give the simulation results for E(*D*) = 0.1 *−* 0.3, respectively, when *Y* is weakly negatively associated with *D*. The corresponding results when *Y* is moderately associated with *D* are given in Tables 1(d)-(f).

We consider three different methods for estimating E(*Y*): Imputation, partial identification bounds without any assumptions (worst case bounds) and partial identification bounds using instrument variables. For the imputation method, we use all the observed variables in the simulation study, i.e., the instruments. For partial identification, we used the worst case bounds that do not make any assumptions, and also the method proposed in this article.

Each combination of parameters corresponds to four rows of results. The first row shows the proportion of times, out of 1000 simulations, the approximate 95 percent confidence intervals include E(*Y*). The second row gives the lower confidence limits, averaged over 1000 simulations. The third row gives the upper confidence limits, averaged over 1000 simulations. The fourth row gives the average width of the confidence intervals. The results are given in Tables S.1 (a)-(f).

**Table S.1:**
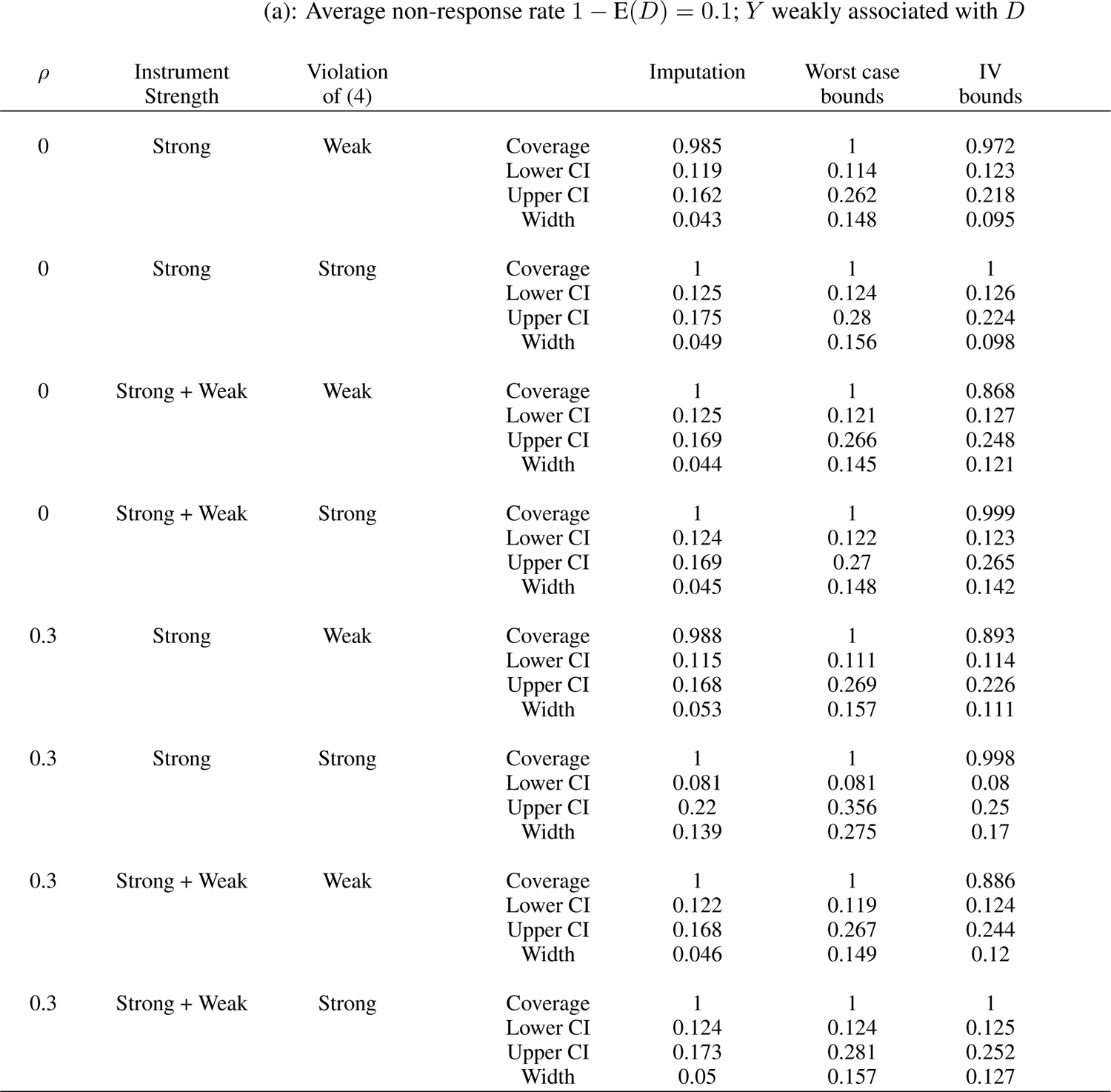
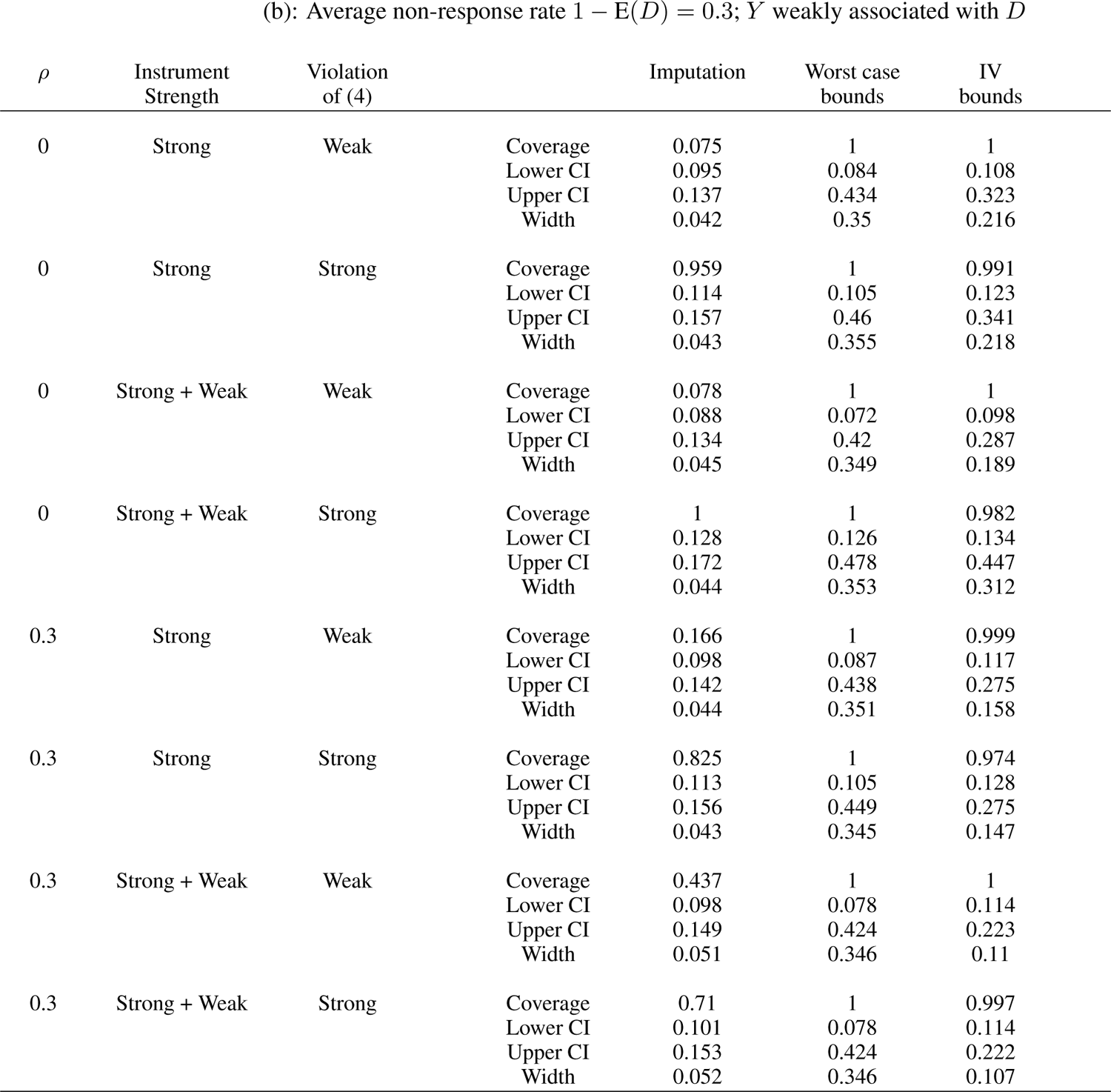
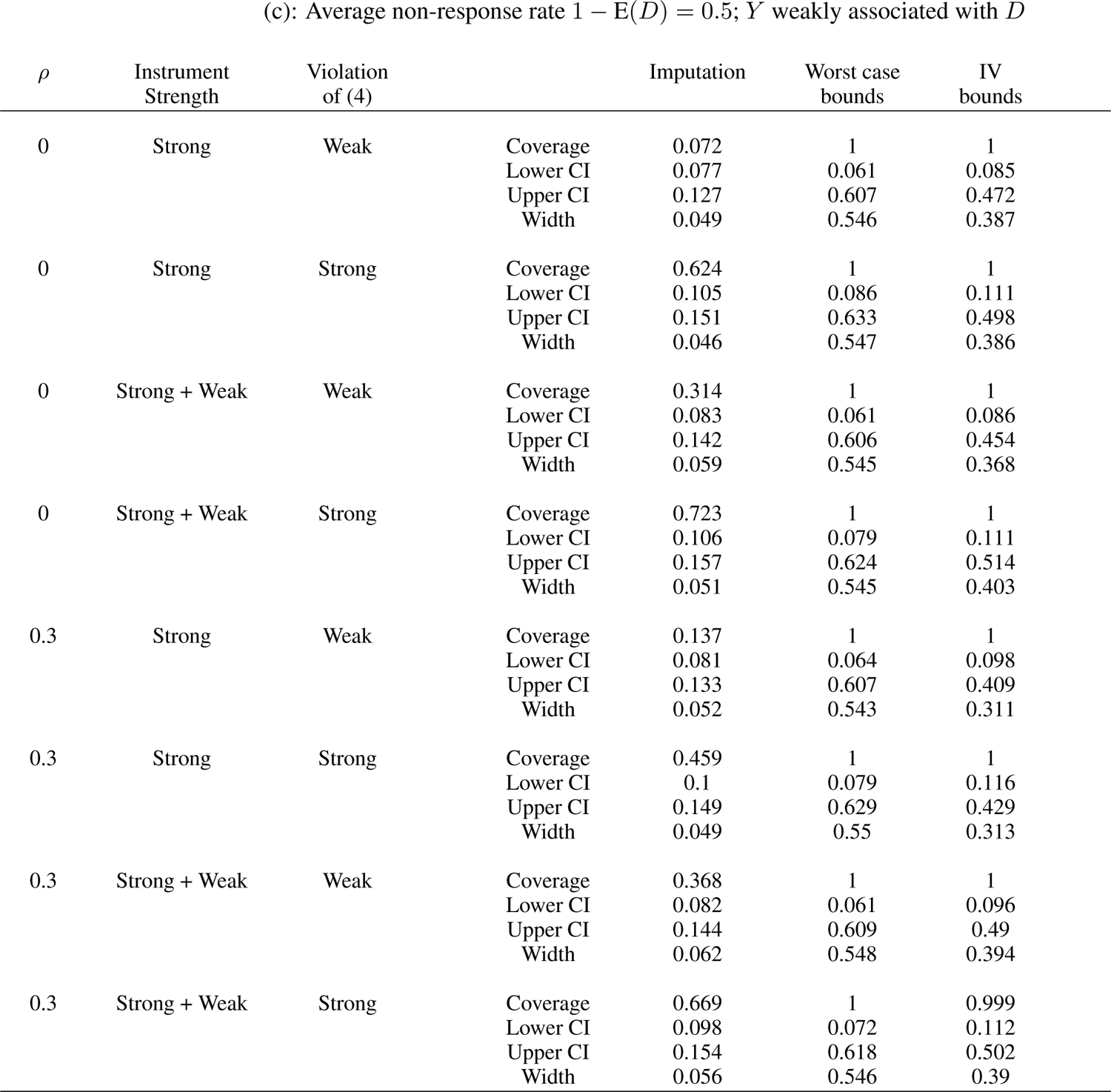
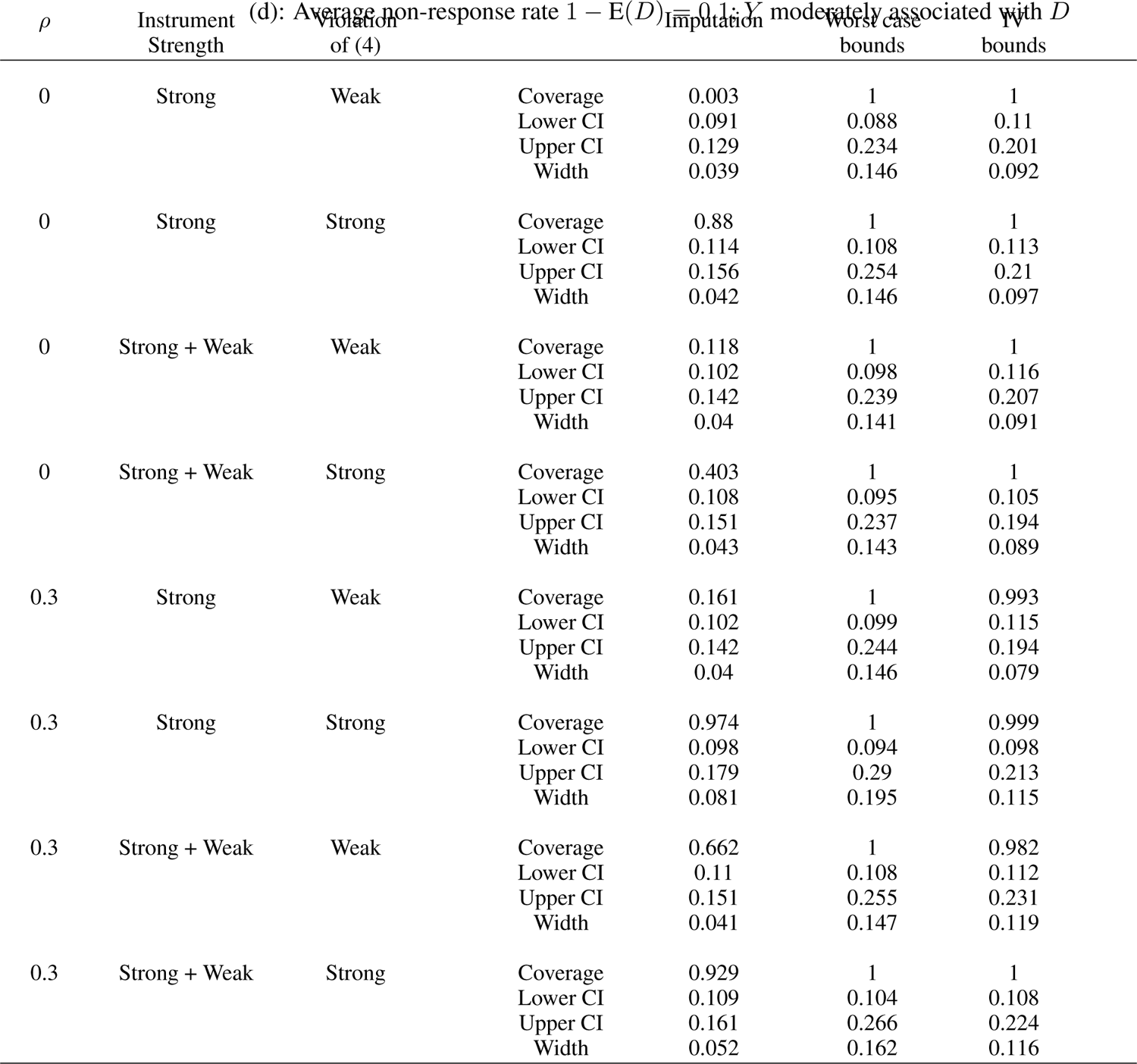
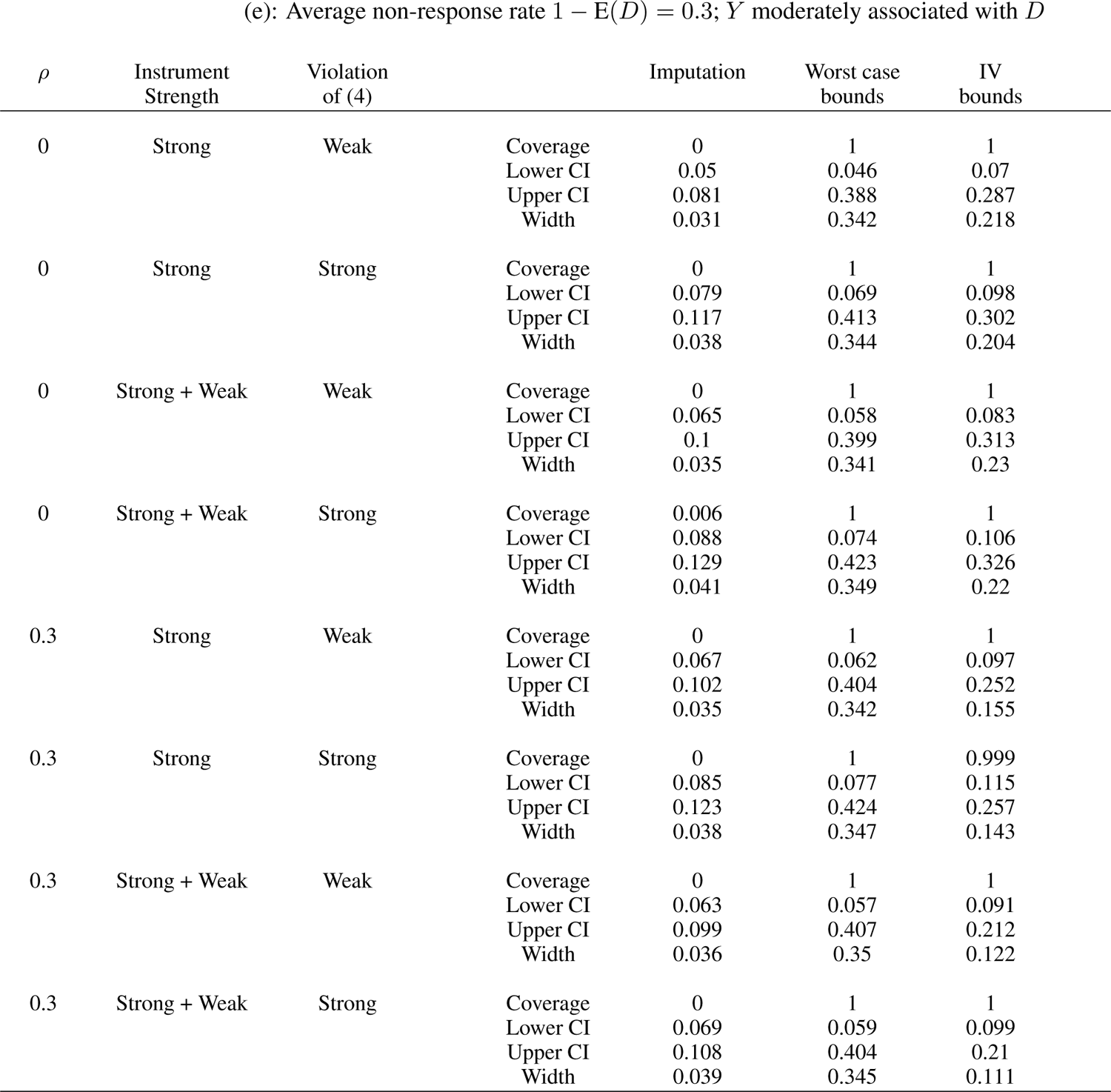
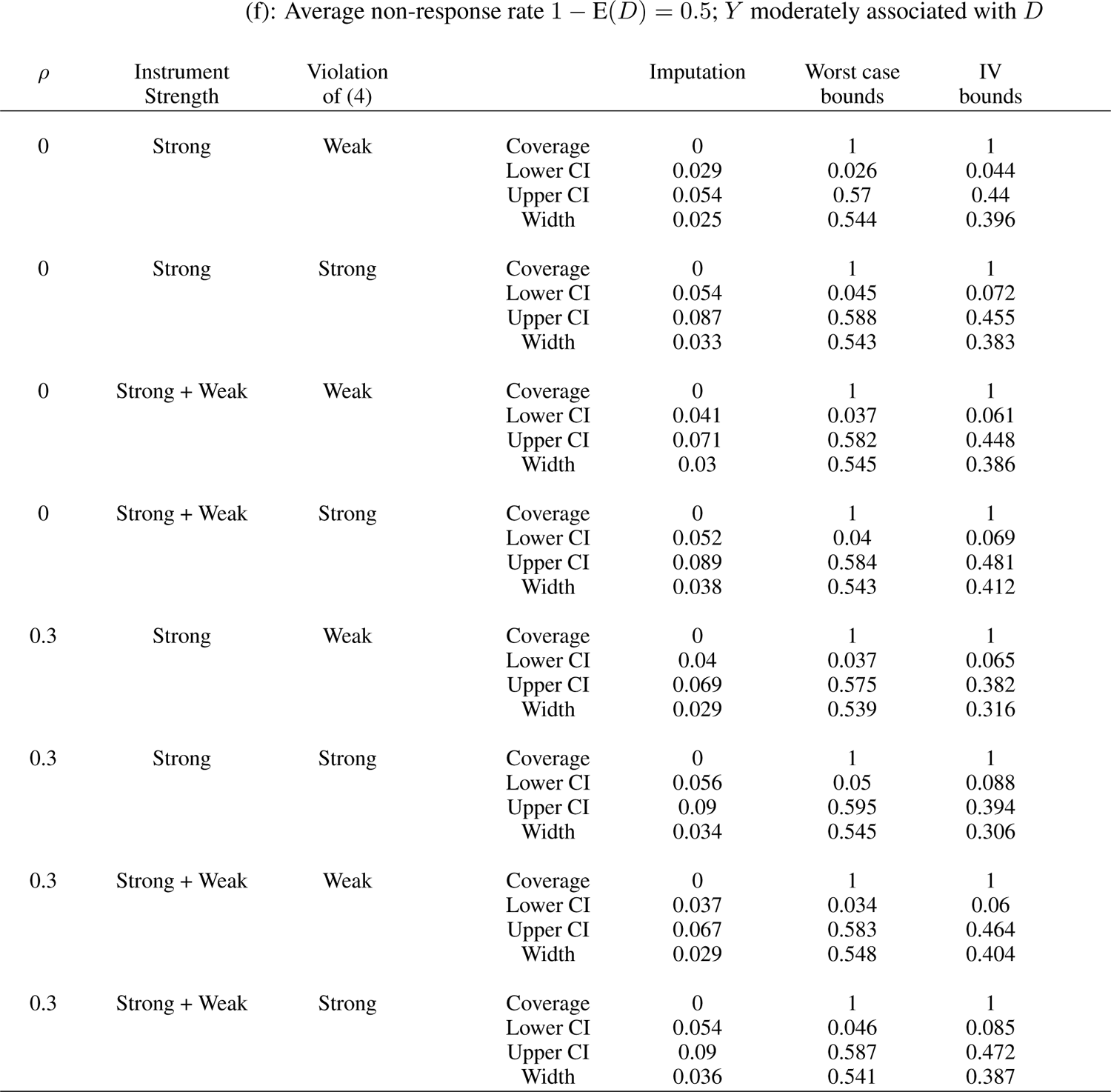
Partial identification of E(*Y*) with *L* = 5 instruments and *s* = 3 valid instruments; E(*Y*) fixed at 0.15; *ρ* = *ρ*_1_ = *ρ*_2_ gives correlation between pairs of valid (invalid) instruments; instruments either all strong or a mixture of strong + weak; the last *L − s* instruments either weakly or strongly violate (4).

### S.4 HIV prevalence using 2007 Zambia DHS

We use the 2007 Zambia DHS data to study non-response adjustment using partial identification bounds. We compare the results to conventional non-response adjustment using imputation. For partial identification, we consider worst case bounds without making any assumptions and also instrumental variable bounds. For the instrumental variable bounds, we use six candidate instrumental variables: iv.lan (whether the language used in the questionnaire or interview is the same as the respondent’s language, yes *vs.* no), iv.firstday (whether the interview was conducted on the first day of the interviews, yes *vs.* no), iv.interviewer (number of interviews the interviewer has performed, *<* 50, 50 *−* 100, 100 *−* 200, *>* 200), iv.mon (whether the interview was carried out during a month of harvest or planting, yes *vs.* no), iv.doa (whether the respondent has known someone who has died of AIDS yes *vs.* no).

The standard non-response adjustment is an imputation analysis on those who are not tested to adjust for potential biases [27]. The individuals in the survey can be classified into one of three groups: (a) those who participated in the household and individual surveys and tested (b) those who participated in the household and individual surveys but not tested and (c) those who only participated in the household surveys. For those in groups (b) and (c), their HIV test results are absent.

For individuals in groups (b) and (c), their probability of HIV is predicted based on multivariate models using data from those who were tested. A logistic regression model is used to calculate HIV probability separately for groups (b) and (c). For group (b), the variables used in the model include the following household survey variables: age, education, wealth quintile, residence, and geographic region, as well as the following variables from the individual survey: marital union, current work status, media exposure, religion, STI or STI symptoms in past 12 months, cigarette smoking/tobacco use, age at first sex, number of sex partners in past 12 months, higher-risk sex in past 12 months, condom use at last sex in past 12 months, and willingness to care for a family member with AIDS. Prediction for group (c) uses only the household variables. The models are used to impute HIV statuses for individuals in groups (b) and (c) and the results are combined with those in group (a) to form adjusted HIV prevalence estimates for the population.

For the partial identification bounds method proposed in this paper, we require instruments. We consider 5 candidate instruments: iv.lan (whether the language used in the questionnaire or interview is the same as the respondent’s language, yes *vs.* no), iv.firstday (whether the interview was conducted on the first day of the interviews, yes *vs.* no), iv.interviewer (number of interviews the interviewer has performed, *<* 50, 50 *−* 100, 100 *−* 200, *>* 200), iv.mon (whether the interview was carried out during a month of harvest or planting, yes *vs.* no), iv.doa (whether the respondent has known someone who has died of AIDS yes *vs.* no). We assume *a* = 3, that is, at least 3 out of the 5 candidates are valid.

We examine HIV prevalence between genders, and across different demographic, socio-economic and behavioural groups (Table S.2). Overall prevalence for women (16.1%) is higher than men (12.3%). In addition, this difference is consistent across all strata groups we examined. There are also significant differences among groups within a strata. For example, women at the lowest wealth quintile has a prevalence of only 8.8% compared to those in the highest two quintiles with over 20% prevalence.

Our results (Table S.3) show that across all scenarios, the imputation method gives very similar results to the unadjusted results. The partial identification bounds always give confidence intervals that are much wider. Between the two partial identification methods, the worst case is always less precise than the method proposed in this paper. The improved precision of the proposed method comes from a big reduction of the upper confidence interval. The improvement ranges from about 10% to 30%.

**Table S.2:** Observed proportions of HIV positive among the tested in 2007 Zambia DHS

|  |  | Women |  | Men |  |
| --- | --- | --- | --- | --- | --- |
|  |  | Observed<br>HIV prevalence | Number | Observed<br>HIV prevalence | Number |
| All |  | 0.161 | 5713 | 0.123 | 5163 |
| Age | 15-19 | 0.058 | 1256 | 0.035 | 1109 |
|  | 20-24 | 0.119 | 1119 | 0.053 | 830 |
|  | 25-29 | 0.198 | 1102 | 0.115 | 772 |
|  | 30-34 | 0.258 | 841 | 0.174 | 746 |
|  | 35-39 | 0.25 | 588 | 0.223 | 594 |
|  | 40-44 | 0.182 | 434 | 0.24 | 390 |
|  | 45-49 | 0.122 | 373 | 0.183 | 318 |
| Religion | Catholic | 0.142 | 1086 | 0.116 | 1080 |
|  | Protestant | 0.167 | 4525 | 0.125 | 3921 |
| Location | Large city | 0.247 | 450 | 0.205 | 439 |
|  | Small city | 0.261 | 424 | 0.159 | 339 |
|  | Town | 0.202 | 1662 | 0.126 | 1410 |
|  | Countryside | 0.11 | 3177 | 0.095 | 2975 |
| Wealth quintile | 1st | 0.088 | 903 | 0.07 | 938 |
|  | 2nd | 0.096 | 987 | 0.098 | 771 |
|  | 3rd | 0.133 | 1122 | 0.104 | 1044 |
|  | 4th | 0.229 | 1387 | 0.18 | 1271 |
|  | 5th | 0.216 | 1314 | 0.138 | 1139 |
| Education | ≤ 6 | 0.123 | 2516 | 0.085 | 1647 |
|  | > 6 | 0.191 | 3197 | 0.14 | 3516 |
| Married | No | 0.183 | 2269 | 0.083 | 2260 |
|  | Yes | 0.147 | 3444 | 0.155 | 2901 |
| Partners last 12m | 0 | 0.153 | 1440 | 0.066 | 1223 |
|  | 1 | 0.161 | 4188 | 0.127 | 3113 |
|  | 1+ | 0.316 | 82 | 0.199 | 815 |
| High risk sex last 12m | No | 0.151 | 4903 | 0.118 | 3683 |
|  | Yes | 0.231 | 807 | 0.136 | 1468 |
| Condom use last sex | No | 0.15 | 5141 | 0.111 | 4260 |
|  | Yes | 0.268 | 569 | 0.179 | 898 |
| STD last 12m | No | 0.153 | 5428 | 0.113 | 4878 |
|  | Yes | 0.339 | 264 | 0.306 | 269 |
| Age first sex | Never | 0.037 | 726 | 0.035 | 674 |
|  | ≤ 15 | 0.173 | 1905 | 0.13 | 1652 |
|  | > 15 | 0.183 | 3080 | 0.141 | 2834 |
| Sex last 12m | No | 0.153 | 1440 | 0.066 | 1223 |
|  | Yes | 0.164 | 4270 | 0.141 | 3928 |
| Ever tested for HIV | No | 0.132 | 3339 | 0.108 | 3945 |
|  | Yes | 0.206 | 2349 | 0.173 | 1216 |

**Table S.3:** 95% confidence intervals for HIV prevalence estimates in 2007 Zambia DHS

|  |  |  | Women |  |  |  | Men |  |  |  |
| --- | --- | --- | --- | --- | --- | --- | --- | --- | --- | --- |
|  |  |  | Unadjusted | Imputation | Worst case bounds | IV bounds | Unadjusted | Imputation | Worst case bounds | IV bounds |
| All |  | LCI | 0.153 | 0.152 | 0.118 | 0.123 | 0.116 | 0.118 | 0.084 | 0.087 |
|  |  | UCI | 0.169 | 0.169 | 0.361 | 0.339 | 0.13 | 0.133 | 0.375 | 0.322 |
|  |  | Width | 0.016 | 0.017 | 0.243 | 0.216 | 0.014 | 0.015 | 0.291 | 0.235 |
| Age | 15-19 | LCI | 0.038 | 0.041 | 0.029 | 0.023 | 0.018 | 0.021 | 0.014 | 0.011 |
|  |  | UCI | 0.078 | 0.073 | 0.305 | 0.274 | 0.051 | 0.045 | 0.32 | 0.278 |
|  |  | Width | 0.04 | 0.032 | 0.276 | 0.251 | 0.033 | 0.024 | 0.306 | 0.267 |
|  | 20-24 | LCI | 0.099 | 0.105 | 0.073 | 0.076 | 0.034 | 0.038 | 0.024 | 0.025 |
|  |  | UCI | 0.139 | 0.136 | 0.36 | 0.331 | 0.073 | 0.067 | 0.352 | 0.317 |
|  |  | Width | 0.04 | 0.031 | 0.287 | 0.255 | 0.039 | 0.029 | 0.328 | 0.292 |
|  | 25-29 | LCI | 0.181 | 0.186 | 0.142 | 0.147 | 0.097 | 0.104 | 0.068 | 0.085 |
|  |  | UCI | 0.216 | 0.213 | 0.392 | 0.383 | 0.132 | 0.129 | 0.406 | 0.358 |
|  |  | Width | 0.035 | 0.027 | 0.25 | 0.236 | 0.035 | 0.025 | 0.338 | 0.273 |
|  | 30-34 | LCI | 0.237 | 0.239 | 0.187 | 0.202 | 0.156 | 0.17 | 0.112 | 0.124 |
|  |  | UCI | 0.279 | 0.272 | 0.44 | 0.401 | 0.192 | 0.197 | 0.425 | 0.386 |
|  |  | Width | 0.042 | 0.033 | 0.253 | 0.199 | 0.036 | 0.027 | 0.313 | 0.262 |
|  | 35-39 | LCI | 0.223 | 0.234 | 0.174 | 0.178 | 0.199 | 0.211 | 0.145 | 0.154 |
|  |  | UCI | 0.278 | 0.276 | 0.443 | 0.408 | 0.246 | 0.246 | 0.459 | 0.378 |
|  |  | Width | 0.055 | 0.042 | 0.269 | 0.23 | 0.047 | 0.035 | 0.314 | 0.224 |
|  | 40-44 | LCI | 0.151 | 0.163 | 0.12 | 0.131 | 0.212 | 0.221 | 0.163 | 0.168 |
|  |  | UCI | 0.212 | 0.211 | 0.391 | 0.342 | 0.268 | 0.262 | 0.455 | 0.413 |
|  |  | Width | 0.061 | 0.048 | 0.271 | 0.211 | 0.056 | 0.041 | 0.292 | 0.245 |
|  | 45-49 | LCI | 0.09 | 0.101 | 0.072 | 0.073 | 0.153 | 0.172 | 0.116 | 0.129 |
|  |  | UCI | 0.155 | 0.152 | 0.333 | 0.315 | 0.211 | 0.214 | 0.431 | 0.382 |
|  |  | Width | 0.065 | 0.051 | 0.261 | 0.242 | 0.058 | 0.042 | 0.315 | 0.253 |
| Religion | Catholic | LCI | 0.122 | 0.132 | 0.097 | 0.105 | 0.097 | 0.106 | 0.079 | 0.081 |
|  |  | UCI | 0.163 | 0.166 | 0.339 | 0.31 | 0.135 | 0.133 | 0.313 | 0.297 |
|  |  | Width | 0.041 | 0.034 | 0.242 | 0.205 | 0.038 | 0.027 | 0.234 | 0.216 |
|  | Protestant | LCI | 0.159 | 0.158 | 0.127 | 0.136 | 0.116 | 0.117 | 0.093 | 0.099 |
|  |  | UCI | 0.175 | 0.171 | 0.341 | 0.325 | 0.134 | 0.13 | 0.315 | 0.303 |
|  |  | Width | 0.016 | 0.013 | 0.214 | 0.189 | 0.018 | 0.013 | 0.222 | 0.204 |

Table S.3: Continued
|  |  |  | Women |  |  |  | Men |  |  |  |
| --- | --- | --- | --- | --- | --- | --- | --- | --- | --- | --- |
|  |  |  | Unadjusted | Imputation | Worst case bounds | IV bounds | Unadjusted | Imputation | Worst case bounds | IV bounds |
| Location | Large city | LCI | 0.224 | 0.227 | 0.155 | 0.163 | 0.177 | 0.185 | 0.115 | 0.114 |
|  |  | UCI | 0.27 | 0.263 | 0.504 | 0.466 | 0.233 | 0.226 | 0.505 | 0.468 |
|  |  | Width | 0.046 | 0.036 | 0.349 | 0.303 | 0.056 | 0.041 | 0.39 | 0.354 |
|  | Small city | LCI | 0.234 | 0.237 | 0.173 | 0.2 | 0.135 | 0.147 | 0.08 | 0.081 |
|  |  | UCI | 0.289 | 0.28 | 0.482 | 0.462 | 0.183 | 0.183 | 0.516 | 0.434 |
|  |  | Width | 0.055 | 0.043 | 0.309 | 0.262 | 0.048 | 0.036 | 0.436 | 0.353 |
|  | Town | LCI | 0.184 | 0.187 | 0.146 | 0.154 | 0.111 | 0.118 | 0.078 | 0.094 |
|  |  | UCI | 0.221 | 0.215 | 0.388 | 0.369 | 0.141 | 0.141 | 0.401 | 0.325 |
|  |  | Width | 0.037 | 0.028 | 0.242 | 0.215 | 0.03 | 0.023 | 0.323 | 0.231 |
|  | Countryside | LCI | 0.1 | 0.099 | 0.077 | 0.081 | 0.084 | 0.085 | 0.064 | 0.072 |
|  |  | UCI | 0.12 | 0.116 | 0.32 | 0.304 | 0.106 | 0.101 | 0.328 | 0.296 |
|  |  | Width | 0.02 | 0.017 | 0.243 | 0.223 | 0.022 | 0.016 | 0.264 | 0.224 |
| Wealth quintile | 1st | LCI | 0.068 | 0.072 | 0.052 | 0.056 | 0.051 | 0.053 | 0.041 | 0.039 |
|  |  | UCI | 0.108 | 0.103 | 0.315 | 0.303 | 0.089 | 0.084 | 0.286 | 0.26 |
|  |  | Width | 0.04 | 0.031 | 0.263 | 0.247 | 0.038 | 0.031 | 0.245 | 0.221 |
|  | 2nd | LCI | 0.078 | 0.082 | 0.06 | 0.059 | 0.077 | 0.08 | 0.056 | 0.073 |
|  |  | UCI | 0.114 | 0.111 | 0.323 | 0.297 | 0.12 | 0.11 | 0.361 | 0.336 |
|  |  | Width | 0.036 | 0.029 | 0.263 | 0.238 | 0.043 | 0.03 | 0.305 | 0.263 |
|  | 3rd | LCI | 0.113 | 0.112 | 0.087 | 0.088 | 0.087 | 0.089 | 0.065 | 0.067 |
|  |  | UCI | 0.153 | 0.144 | 0.351 | 0.315 | 0.121 | 0.115 | 0.364 | 0.321 |
|  |  | Width | 0.04 | 0.032 | 0.264 | 0.227 | 0.034 | 0.026 | 0.299 | 0.254 |
|  | 4th | LCI | 0.213 | 0.215 | 0.165 | 0.176 | 0.164 | 0.167 | 0.115 | 0.113 |
|  |  | UCI | 0.245 | 0.239 | 0.423 | 0.385 | 0.196 | 0.19 | 0.445 | 0.386 |
|  |  | Width | 0.032 | 0.024 | 0.258 | 0.209 | 0.032 | 0.023 | 0.33 | 0.273 |
|  | 5th | LCI | 0.199 | 0.204 | 0.153 | 0.161 | 0.122 | 0.134 | 0.083 | 0.095 |
|  |  | UCI | 0.233 | 0.232 | 0.416 | 0.393 | 0.154 | 0.158 | 0.435 | 0.354 |
|  |  | Width | 0.034 | 0.028 | 0.263 | 0.232 | 0.032 | 0.024 | 0.352 | 0.259 |
| Education | $\leq 6$ | LCI | 0.112 | 0.113 | 0.084 | 0.086 | 0.071 | 0.074 | 0.052 | 0.055 |
|  |  | UCI | 0.134 | 0.132 | 0.349 | 0.321 | 0.098 | 0.096 | 0.345 | 0.306 |
|  |  | Width | 0.022 | 0.019 | 0.265 | 0.235 | 0.027 | 0.022 | 0.293 | 0.251 |
| | $> 6$ | LCI | 0.179 | 0.182 | 0.14 | 0.146 | 0.13 | 0.136 | 0.094 | 0.096 |
|  |  | UCI | 0.203 | 0.202 | 0.379 | 0.354 | 0.15 | 0.152 | 0.394 | 0.337 |
|  |  | Width | 0.024 | 0.02 | 0.239 | 0.208 | 0.02 | 0.016 | 0.3 | 0.241 |

Table S.3: Continued
|  |  |  | Women |  |  |  | Men |  |  |  |
| --- | --- | --- | --- | --- | --- | --- | --- | --- | --- | --- |
|  |  |  | Unadjusted | Imputation | Worst case bounds | IV bounds | Unadjusted | Imputation | Worst case bounds | IV bounds |
| Married | No | LCI | 0.171 | 0.171 | 0.138 | 0.141 | 0.071 | 0.074 | 0.057 | 0.059 |
|  |  | UCI | 0.195 | 0.192 | 0.359 | 0.345 | 0.094 | 0.091 | 0.29 | 0.278 |
|  |  | Width | 0.024 | 0.021 | 0.221 | 0.204 | 0.023 | 0.017 | 0.233 | 0.219 |
|  | Yes | LCI | 0.136 | 0.138 | 0.108 | 0.119 | 0.143 | 0.147 | 0.116 | 0.12 |
|  |  | UCI | 0.158 | 0.157 | 0.33 | 0.3 | 0.167 | 0.164 | 0.337 | 0.32 |
|  |  | Width | 0.022 | 0.019 | 0.222 | 0.181 | 0.024 | 0.017 | 0.221 | 0.2 |
| Partners last 12m | 0 | LCI | 0.134 | 0.137 | 0.106 | 0.111 | 0.047 | 0.052 | 0.037 | 0.037 |
|  |  | UCI | 0.172 | 0.166 | 0.348 | 0.341 | 0.086 | 0.08 | 0.305 | 0.282 |
|  |  | Width | 0.038 | 0.029 | 0.242 | 0.23 | 0.039 | 0.028 | 0.268 | 0.245 |
|  | 1 | LCI | 0.151 | 0.153 | 0.122 | 0.128 | 0.116 | 0.12 | 0.093 | 0.099 |
|  |  | UCI | 0.17 | 0.169 | 0.338 | 0.311 | 0.138 | 0.138 | 0.319 | 0.302 |
|  |  | Width | 0.019 | 0.016 | 0.216 | 0.183 | 0.022 | 0.018 | 0.226 | 0.203 |
|  | 1+ | LCI | 0.254 | 0.27 | 0.228 | 0.264 | 0.174 | 0.181 | 0.15 | 0.159 |
|  |  | UCI | 0.379 | 0.358 | 0.479 | 0.525 | 0.224 | 0.217 | 0.351 | 0.33 |
|  |  | Width | 0.125 | 0.088 | 0.251 | 0.261 | 0.05 | 0.036 | 0.201 | 0.171 |
| High risk sex last 12m | No | LCI | 0.142 | 0.143 | 0.113 | 0.119 | 0.108 | 0.11 | 0.085 | 0.087 |
|  |  | UCI | 0.159 | 0.158 | 0.332 | 0.313 | 0.127 | 0.126 | 0.32 | 0.31 |
|  |  | Width | 0.017 | 0.015 | 0.219 | 0.194 | 0.019 | 0.016 | 0.235 | 0.223 |
|  | Yes | LCI | 0.206 | 0.211 | 0.169 | 0.171 | 0.119 | 0.122 | 0.101 | 0.103 |
|  |  | UCI | 0.255 | 0.248 | 0.404 | 0.384 | 0.154 | 0.148 | 0.307 | 0.286 |
|  |  | Width | 0.049 | 0.037 | 0.235 | 0.213 | 0.035 | 0.026 | 0.206 | 0.183 |
| Condom use last sex | No | LCI | 0.142 | 0.142 | 0.113 | 0.118 | 0.104 | 0.105 | 0.082 | 0.088 |
|  |  | UCI | 0.159 | 0.157 | 0.332 | 0.315 | 0.119 | 0.118 | 0.306 | 0.291 |
|  |  | Width | 0.017 | 0.015 | 0.219 | 0.197 | 0.015 | 0.013 | 0.224 | 0.203 |
|  | Yes | LCI | 0.245 | 0.252 | 0.202 | 0.204 | 0.162 | 0.166 | 0.133 | 0.137 |
|  |  | UCI | 0.29 | 0.288 | 0.421 | 0.39 | 0.196 | 0.191 | 0.358 | 0.332 |
|  |  | Width | 0.045 | 0.036 | 0.219 | 0.186 | 0.034 | 0.025 | 0.225 | 0.195 |
| STD last 12m | No | LCI | 0.145 | 0.146 | 0.116 | 0.122 | 0.105 | 0.107 | 0.083 | 0.089 |
|  |  | UCI | 0.161 | 0.16 | 0.333 | 0.313 | 0.121 | 0.12 | 0.307 | 0.289 |
|  |  | Width | 0.016 | 0.014 | 0.217 | 0.191 | 0.016 | 0.013 | 0.224 | 0.2 |
|  | Yes | LCI | 0.304 | 0.312 | 0.26 | 0.283 | 0.272 | 0.281 | 0.235 | 0.238 |
|  |  | UCI | 0.373 | 0.366 | 0.475 | 0.422 | 0.34 | 0.329 | 0.453 | 0.447 |
|  |  | Width | 0.069 | 0.054 | 0.215 | 0.139 | 0.068 | 0.048 | 0.218 | 0.209 |

Table S.3: Continued
|  |  |  | Women |  |  |  | Men |  |  |  |
| --- | --- | --- | --- | --- | --- | --- | --- | --- | --- | --- |
|  |  |  | Unadjusted | Imputation | Worst case bounds | IV bounds | Unadjusted | Imputation | Worst case bounds | IV bounds |
| Age first sex | Never | LCI | 0.016 | 0.021 | 0.012 | 0.015 | 0.014 | 0.018 | 0.011 | 0.009 |
|  |  | UCI | 0.057 | 0.053 | 0.271 | 0.267 | 0.057 | 0.052 | 0.31 | 0.298 |
|  |  | Width | 0.041 | 0.032 | 0.259 | 0.252 | 0.043 | 0.034 | 0.299 | 0.289 |
|  | ≤ 15 | LCI | 0.161 | 0.161 | 0.132 | 0.139 | 0.117 | 0.121 | 0.097 | 0.097 |
|  |  | UCI | 0.185 | 0.181 | 0.336 | 0.315 | 0.144 | 0.141 | 0.302 | 0.298 |
|  |  | Width | 0.024 | 0.02 | 0.204 | 0.176 | 0.027 | 0.02 | 0.205 | 0.201 |
|  | > 15 | LCI | 0.172 | 0.175 | 0.136 | 0.144 | 0.131 | 0.135 | 0.105 | 0.113 |
|  |  | UCI | 0.194 | 0.192 | 0.364 | 0.34 | 0.151 | 0.15 | 0.33 | 0.313 |
|  |  | Width | 0.022 | 0.017 | 0.228 | 0.196 | 0.02 | 0.015 | 0.225 | 0.2 |
| Sex last 12m | No | LCI | 0.134 | 0.137 | 0.106 | 0.111 | 0.047 | 0.052 | 0.037 | 0.037 |
|  |  | UCI | 0.172 | 0.166 | 0.348 | 0.341 | 0.086 | 0.08 | 0.305 | 0.282 |
|  |  | Width | 0.038 | 0.029 | 0.242 | 0.23 | 0.039 | 0.028 | 0.268 | 0.245 |
|  | Yes | LCI | 0.154 | 0.156 | 0.124 | 0.133 | 0.132 | 0.135 | 0.107 | 0.114 |
|  |  | UCI | 0.173 | 0.171 | 0.339 | 0.311 | 0.151 | 0.15 | 0.322 | 0.302 |
|  |  | Width | 0.019 | 0.015 | 0.215 | 0.178 | 0.019 | 0.015 | 0.215 | 0.188 |
| Ever tested for HIV | No | LCI | 0.12 | 0.121 | 0.096 | 0.1 | 0.099 | 0.101 | 0.079 | 0.084 |
|  |  | UCI | 0.144 | 0.14 | 0.323 | 0.3 | 0.117 | 0.116 | 0.305 | 0.289 |
|  |  | Width | 0.024 | 0.019 | 0.227 | 0.2 | 0.018 | 0.015 | 0.226 | 0.205 |
|  | Yes | LCI | 0.191 | 0.196 | 0.155 | 0.169 | 0.157 | 0.162 | 0.128 | 0.136 |
|  |  | UCI | 0.221 | 0.219 | 0.373 | 0.361 | 0.19 | 0.186 | 0.354 | 0.351 |
|  |  | Width | 0.03 | 0.023 | 0.218 | 0.192 | 0.033 | 0.024 | 0.226 | 0.215 |

## Footnotes

† Our expressions differ from those in equations (6) and (7) of [22] by a factor of √*n* because they use the notation of *σ-·*/√n to denote standard error.

## Notes

### Competing Interest Statement

The authors have declared no competing interest.

### Funding Statement

No funding

